# Optimally Pooled Viral Testing

**DOI:** 10.1101/2020.07.05.20145805

**Authors:** Dor Ben-Amotz

**Affiliations:** Purdue University, Department of Chemistry, West Lafayette, IN 47907

## Abstract

It has long been known that pooling samples may be used to minimize the total number of tests required in order to identify each infected individual in a population. Pooling is most advantageous in populations with low infection (positivity) rates, but is expected to remain better than non-pooled testing in populations with infection rates up to 30%. For populations with infection rates lower than 10%, additional testing efficiency may be realized by performing a second round of pooling to test all the samples in the positive first-round pools. The present predictions are validated by recent COVID-19 (SARS-CoV-2) pooled testing and detection sensitivity measurements performed using non-optimal pool sizes, and quantify the additional improvement in testing efficiency that could have been obtained using optimal pooling. Although large pools are most advantageous for testing populations with very low infection rates, they are predicted to become highly non-optimal with increasing infection rate, while pool sizes smaller than 10 remain near-optimal over a broader range of infection rates.

## Background

The advantages of pooled testing in applications ranging from disease screening to manufacturing quality assurance have long been appreciated.^1^ Efficiently and practically containing viral outbreaks requires minimizing the total number of tests needed in order to uniquely identify every positive individual. This may be achieved using pooled testing, given a sufficiently sensitive diagnostic test with an acceptably low false-negative detection probability. When applicable, pooled testing a population consisting of a large number *N* of individuals can be achieved with significantly fewer than *N* tests, by initially screening pools containing a mixture of *n* samples, followed by further testing of only the positive pools to uniquely identify each infected individual. The latter tests may either be carried out by separately testing all individuals in the positive first-round pools or by using a second round of pooling to more efficiently identify each infected individuals in the positive first-round pools. The present results provide predictions regarding optimal first and second round pool sizes for a population with an infection probabilities of 0.001 ≤ *p* < 0.3 (corresponding to an infection rates of 0.1% to 30%), as well as the range of infection rates over which a given fixed pool size remains near-optimal.

The primary aim of this work is to provide SARS-CoV-2 (COVID-19) testing centers with practical guidance regarding the efficient implementation of pooled testing. Specifically, the present predictions (which are most conveniently summarized in Table 1) may be used to aid both in the selection an optimal pool size for a population with a given estimated infection rate, as well as in the real time adjustment of the pool size to better match the actual evolving infection rate obtained from day to day measurements. Most importantly, the present results facilitate significantly expanding the number of individuals that can be tests using a given number of tests, using either one or two rounds of pooled testing.

**Table 1:**
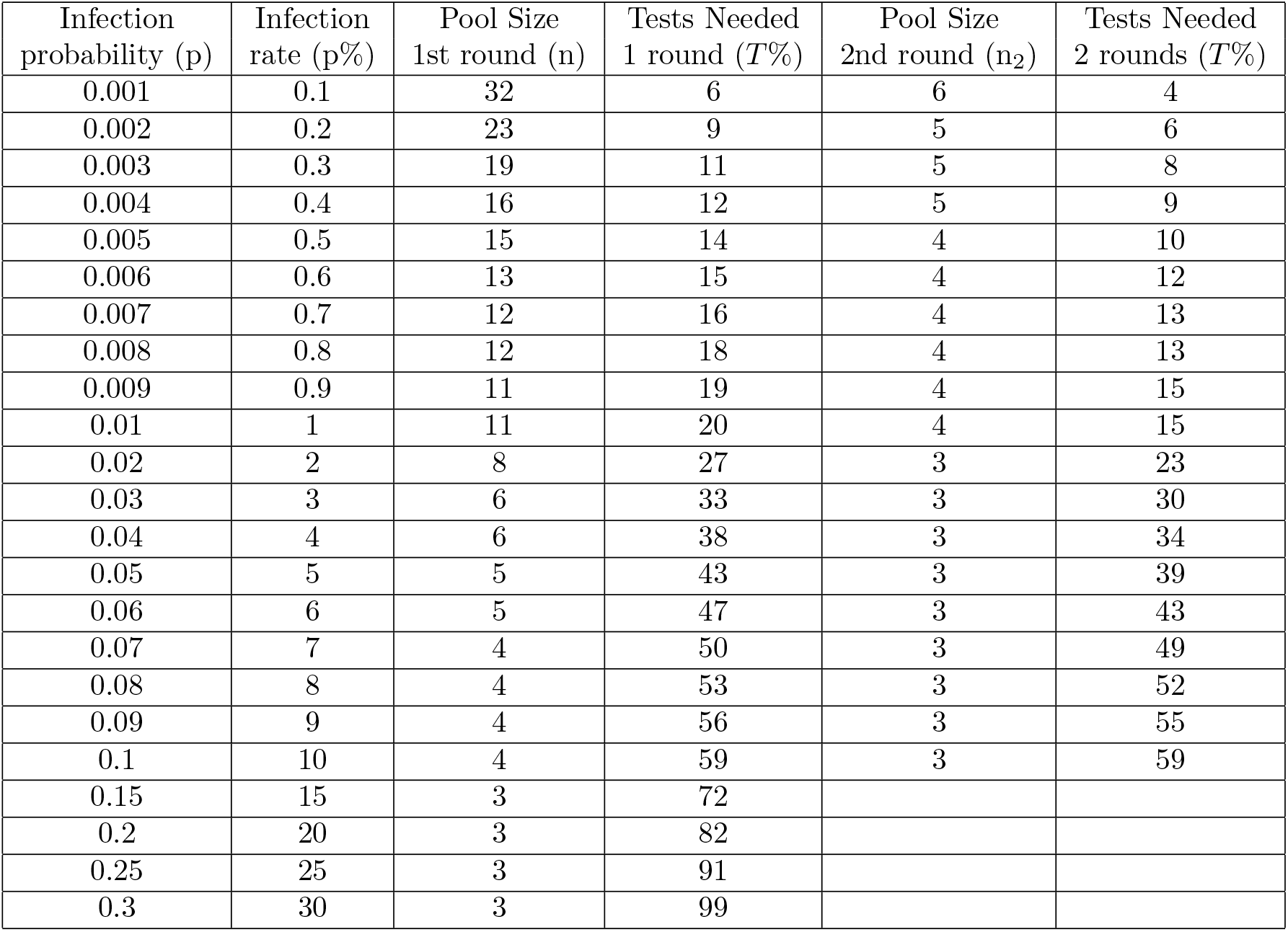
Optimally Pooled Testing Predictions.

The present predictions are obtained assuming that the population of interest has a uniform infection rate, which may be detected perfect accuracy and specificity. In spite of these idealizations, the practical utility of the predictions are quantitatively validated by recent SARS-CoV-2 pooled testing data.^2–6^ The results indicate that pooled testing can significantly reduce the number of the SARS-CoV-2 tests required to identify each positive individual, even in populations with infection rates above 10%, although pooled testing is most advantageous in populations with lower infection rates. The field testing validation studies also confirm that the present predictions provide conservative testing efficiency estimates, as non-uniform clustering of infections is predicted to lead to an increase in testing efficiency, above that predicted assuming a uniform infection rate.

The optimal pool size, *n*, for a population with a given infection probability *p* was first obtained obtained by Dorfman^1^ (and has since spawned numerous generalizations). ^5,7,8^ Here Dorfman’s results are extended to yield practically useful predictions of the range of infection rates over which a given fixed pool size remains nearly optimal, as well as the significant additional efficiency that may be obtainable from using a second round of pooling for populations with 0.001 ≤ *p <* 0.1 (0.1% ≤ *p*% < 10%). For a populations with a very low infection rate of 0.1%, the predicted 1st round optimal pool size is 32. The practicality of using pools this large has recently been demonstrated by showing that a standard RT-qPCR test for SARS-CoV-2 may be used to detect a single positive individual in pools as large as 32, with with 90% accuracy (corresponding to a false negative rate of 10%),^6^ which is consistent with results reported in a recent article in *Lancet*.^2^ However, it is also important to note that tests performed using such large pools are only predicted to be beneficial for populations with a very low (and narrow) range of infection rates, and become highly non-optimal for populations with infection rates exceeding 1%. Thus, for example, for infection rate near or exceeding 10%, optimal pooling efficiency requires using pools smaller than 5.

In practice, these predictions may be used by initially choosing a pool size optimal for the estimate infection rate in the population of interest, and subsequently adjusting the pool size to better match the actual infection probability. These predictions are expected to be most useful in facilitating large scale screening and continuous testing of populations with low infection probabilities for early detection of SARS-CoV-2 outbreaks, to enhance both public safety and economic productivity.

## Methods

The binomial distribution yields the following expression for the probability that there will be *k* infected individuals in a pool of sized *n*, drawn from a population with an infection probability of *p*.^9^

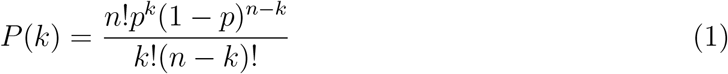

When *k* = 0 this reduces to the following expression for the fraction of pools that are expected to contain no infected individuals, in keeping with Dorfman’s original predictions.^1^

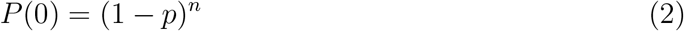

This yields the following expression for the total number of tests *N*_*tests*_ required in order to identify every positive individual in a population of size *N*, when using a pool size of *n*.

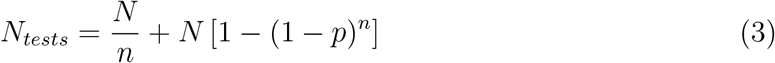

Thus, the average percentage of tests that must be performed in order to identify every infected individual in a population of size *N* is *T* % = 100 × (*N*_*tests*_*/N*). In other words, *T* % represents the average number of tests required to identify each infected individual in a population of size 100, or equivalently *T* %× 1,000 is the number of tests required to do so in a population of 100,000.

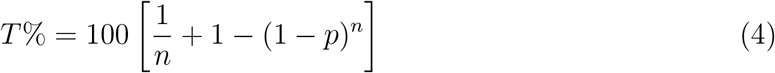

The optimal value of *n* is that which minimizes *T* %, and thus may be obtained by finding the roots of the following expression for the partial derivative of *T* % with respect to *n*, pertaining to a given value of *p*.

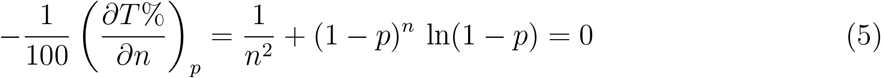

The above expression may be solved numerically using Newton’s method. Alternatively, the optimal pool size may also be obtained iteratively, using an initial guess for the pool size *n*_0_, inserted into the right-hand-side of the following expression, to obtain a better estimate of *n* (where the “Round” operation rounds the result to the nearest positive integer).

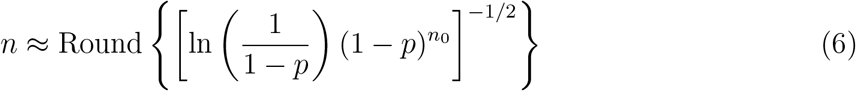

If *n*_0_ is not very similar to *n*, then one may set *n*_0_ = *n* and repeat the process to obtain a better estimate of *n*. This iterative procedure typically converges within a few cycles (whose convergence can be most accurately quantified by removing the Round operation from the right-hand-side of Eq. 6). Note that the optimal pool size may also be quite accurately approximated using 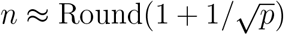.^10^

The infected individuals in the positive first-round pools may in some cases be more efficiently determined using a second round of pooled testing. The average infection probability *p*_2_ in all the positive first round pools is higher than that in the original population because all the non-infected individuals in the negative first round pools have been removed from the population of second round test samples. Thus, the optimal second round pool size *n*_2_ is obtained as follows, where *n*_2_ is the optimal pool size pertaining to an infection probability of *p*_2_ (obtained using Eqs. 5 or 6).

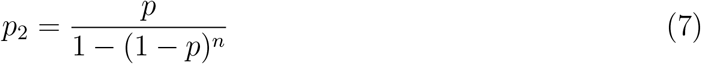

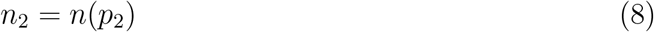

The above results imply that using two rounds of pooled testing is only advantageous for populations with with *p* ≤ 0.1 (as the predicted value of *p*_2_ exceeds 0.3 at higher infection rates). Thus, the following equation predicts the total number of tests required to identify every infected individual when using two rounds of optimal pooling.

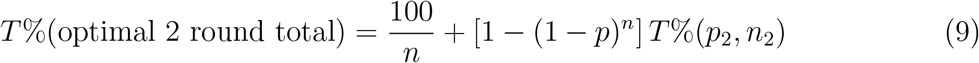

Note that 100*/n* is the number of pools that were tested in the first round (expressed as a percent of total number of tested individuals *N*), and 1 − (1 − *p*)^*n*^ is the fraction of positive first round pools, and thus [1 − (1 − *p*)^*n*^] *T* %(*p*_2_, *n*_2_) is the number of tests required to identify all the positive individuals in those pools, where *T* %(*p*_2_, *n*_2_) is obtained using Eq. 4 (with *p* = *p*_2_ and *n* = *n*_2_).

More generally, Eqs. 4 and 9 may also be used to obtain predictions pertaining the efficiency of non-optimally pooled test, for a population with a given average infection probability *p*, and any chosen values of *n* and *n*_2_.

## Results

Table 1 shows the optimal pooled testing predictions pertaining to populations with uniform infection rates ranging from 0.1% to 30%. The 3rd column contains the predicted optima first-round pool size *n* (obtained using Eqs. 5 or 6), and the 4th column contains the resulting first round *T* % predictions. Note that these values correspond to averages over large populations. For example, for a population with an infection probability of *p* = 0.001 (0.1%), the optimal pool size is 32 and the vast majority of such pools will contain no infected individuals. More specifically, any such pool of size 32 is predicted to have no infections 97% of the time, and the remaining 3% of the pools of size 32 are predicted to contain only one infected individual. The last two columns in Table 1 pertain to predictions obtained when using a second round of pooling to test all the positive first-round pools, as further discussed below.

Figure 1 contains more detailed predictions pertaining to the average number of infected individuals in pools of optimal size, when the overall infection probability ranges from 1% to 30%. Note that at (and below) an infection probability of 1%, essentially all of the positive pools are predicted to contain only one infected individual. At higher infection probabilities a non-negligible number of positive pools are predicted to contain more than one infected individual, but nevertheless most positive pools are predicted to contain only one infected individual. For example, even in a population with an infection probability of 30%, about 34% of the pools of size 3 are predicted to contain no infected individuals, while 45% contain one, and only 21% contain more than one infected individual. However, at this high rate of infection pool testing is no longer adventageous, relative to exhaustively testing every single individual, as indicated by the 4th column in Table 1, which indicates that an average of 99 tests would have to be performed when optimally pool testing a population of 100 individuals that has an infection rate of 30%.

**Figure 1:**
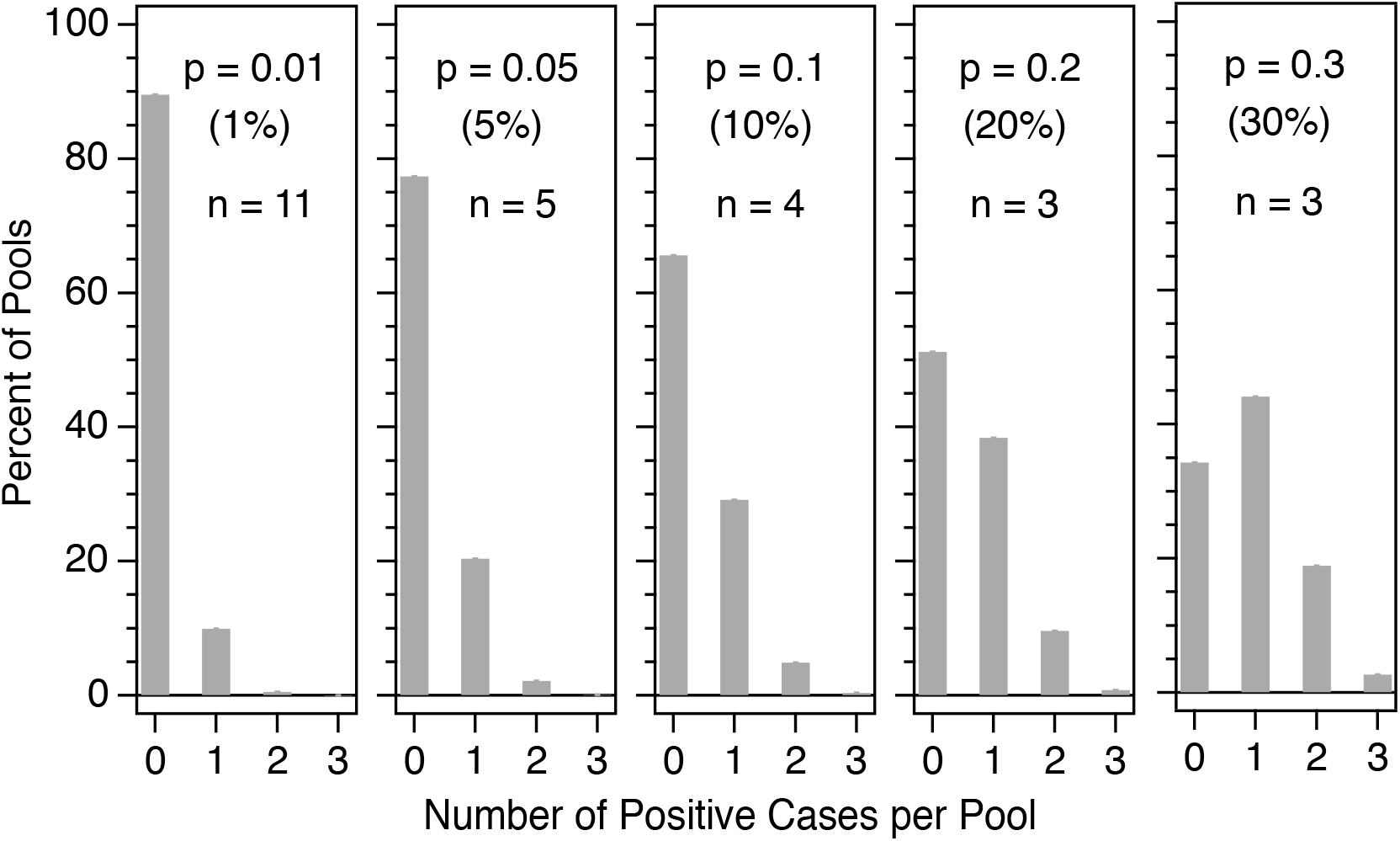
Predicted number of infected individuals in optimally sized pools obtained from populations with average infection rates ranging from 1% to 30%.

Figures 2 and 3 contain graphical predictions pertaining to tests performed using either one or two rounds of optimal pooling, respectively. Figure 2 shows the resulting optimal first round pool size *n* (**a**) and testing percentage T% (**b**) predictions. The insert panels in each figure contain an expanded view of the predictions pertaining to populations with infection rates less than 1% (*p* ≤ 0.01), and the solid curves are optimal pooled testing predictions. The optimal pool size values shown in Table 1 are obtained by rounding the graphical results to the nearest positive integer. The dotted curves in Figure 2**b** show the testing efficiency predictions obtained when using various fixed pool size estimates *n*_0_, indicating that nearoptimal pool sizes produce results that are essentially the same as those obtained using an optimal pool size. More specifically, these predictions indicate that pool sizes of 5, 6, and 7 are expected to produce nearly optimal testing efficiency in populations with average infection rates of 2%-12%, 1%-8%, and 0.7%-6%, respectively (as determined by requiring that T% remain within 3% of its optimal value). The dotted curves in the inset panel in Fig. 2**b** illustrates the fact that large pool sizes are only expected to be optimal over a very narrow range of infection probabilities, and to ra1p0i0dly become significantly non-optimal with increasing infection probability.

**Figure 2:**
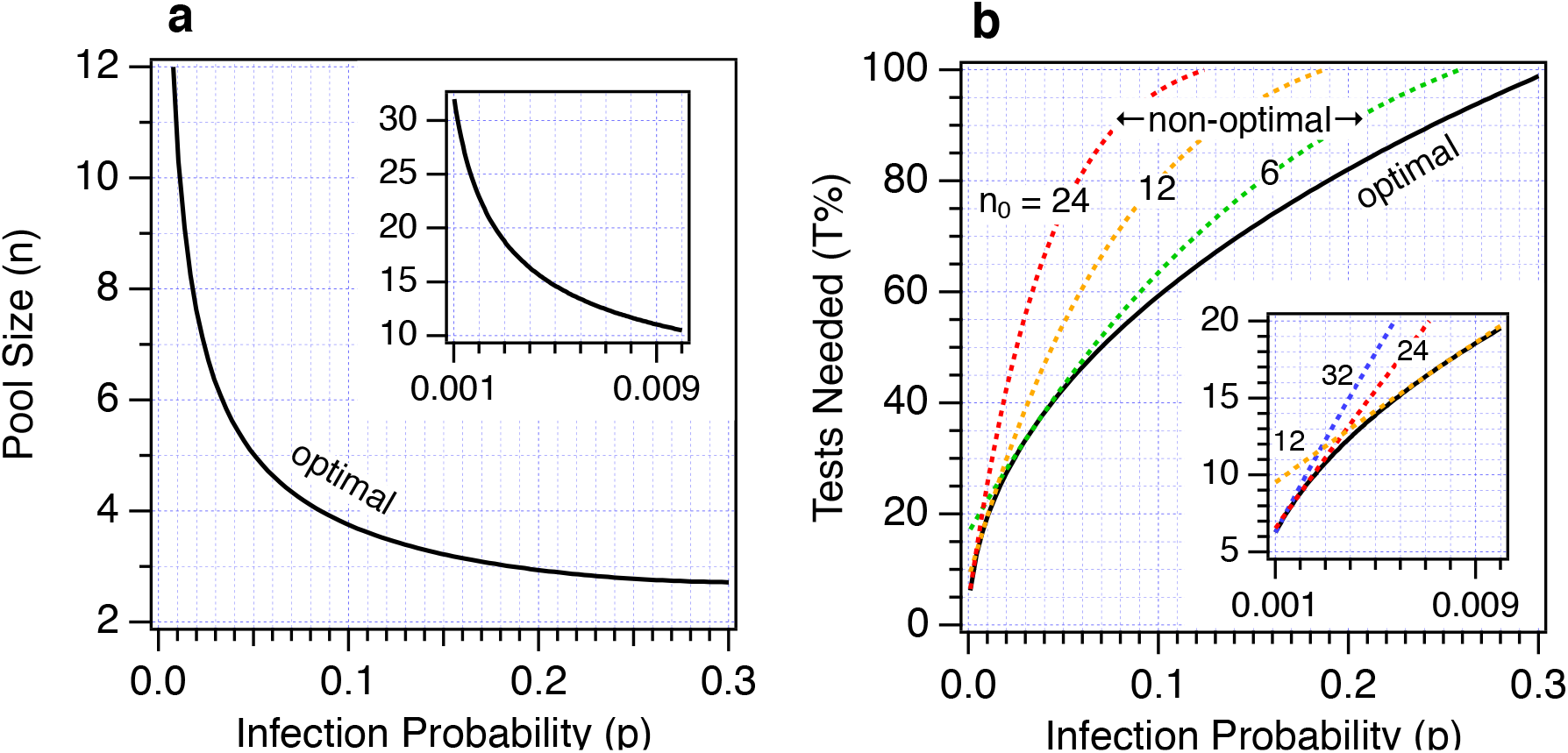
Optimal pool size **a** and testing percentage **b** predictions obtained when applying a single round of pooled testing. The dashed lines in **b** represent the testing percentages obtained using three different fixed pool sizes.

**Figure 3:**
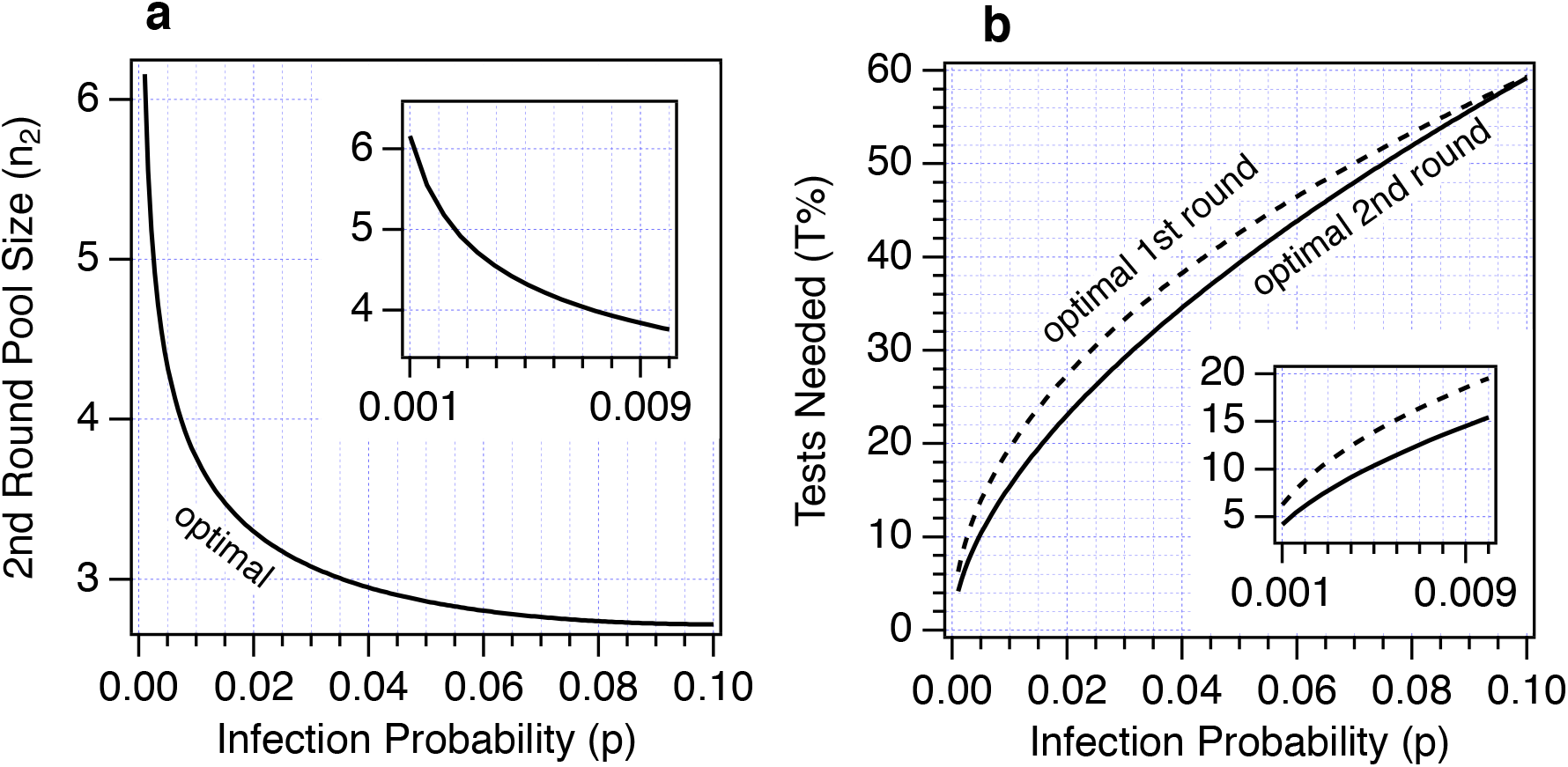
Predicted optimal pool sizes **a** and testing percentages **b** obtained when applying two rounds of pooled testing. The dashed and solid curves in **b** compare the predicted testing efficiencies obtainable using one or two rounds of optimized pool testing, respectively.

Figure 3, as well as the last two columns in Table 1, contain predictions pertaining to the use of two rounds of optimal pooling, performed on the same population of test samples. Specifically, the 5th column in Table 1 contains the optimal second round pool size and the last column contains the predicted average number of tests required to determine all the infected individuals in a population of size 100 when using two rounds of optimal pooling. The second round of optimal pooling is performed by limiting the second round tests to individuals in the positive first round pools, as further described below.

The solid curves in Figure 3 are optimal second round pool testing predictions. Figure 3**a** shows the predicted optimal pool size, *n*_2_, that should be used in order to efficiently re-test all the samples from the positive first round pools. The dashed and solid curves in Figure 3**b**, as well as the 4th and 6th columns in Table 1, compare the first and second round optimal testing efficiency predictions. Note that using a second round of pooling is only predicted to be advantageous for populations with infection rates above 10%. For example, in a population with an infection rate of 1%, one round of pooling is predicted to require an average of 20 tests per 100 individuals, while two rounds of pooling reduces that to 15 tests per 100 individuals. The fractional gain in testing efficiency increases as the infection rate decreases. For example, the required number of tests decreases from 6 to 4 per 100 individuals in a population with an infection rate of 0.1%. Again, note that these predictions represent the average number of tests required per 100 individuals, and the value of 4 arises because approximately 97% of the first round pools of size 32 drawn from such a population are predicted to contain no infected individuals.

Although no prior studies have reported SARS-CoV-2 test results performed using either one or two rounds of optimal pooled testing, the present predictions may nevertheless be critically tested and validated using comparisons with recently reported SARS-CoV-2 pooled test data obtained using non-optimal pool sizes. For example, as recently reported in *Lancet*,^2^ two rounds of pooled tests for SARS-CoV-2 were performed on 1191 samples with an average infection rate of 1.93%, using 1st and 2nd round pool sizes of 30 and 10, respectively. A total of 267 tests were required in order to identify each of the 23 infected individuals in that population, corresponding to *T* % = 100 × (267*/*1191) = 22.4%, which is quite close to the predicted value of *T* % = 23.7% (corresponding to 282 tests) obtained using Eq. 9 assuming the same infection rate and pool sizes. Moreover, the present predictions imply that about 15 fewer tests would have been required if more nearly optimal 1st and 2nd round pool sizes of 8 and 3, respectively, had been used to test the same population.

As another example, an early SARS-CoV-2 pool testing study used a pool size of 10 to test 2888 samples obtained from a population with an average infection rate of 0.07%. ^4^ These pooled tests correctly identified the two infected individuals in this population using a total of 312 tests, or T% = 10.8%, which compares very well with T%=10.7% predicted using Eq. 4. Yet another validation of the present predictions is obtained from recent test of 2160 samples from a population with an infection rate of 0.23% using a pool size of 8,^5^ in which SARS-CoV-2 five infected individuals were identified using at total of 311 tests, or T% = 14.4%, which compares very favorably with T% = 14.3% predicted using Eq. 4. In both of the above examples the present predictions imply that a substantial additional gain in pooled testing efficiency could have been obtained by using a more nearly optimal pooling strategy. Specifically, if the first population ^4^ were tested using optimal 1st and 2nd round pool sizes of 38 and 7, respectively, the required number of test is predicted to decrease *T* % to 3% (from ∼11%), and in the second population^5^ the use of optimal 1st and 2nd round pool sizes of 21 and 5, respectively, is predicted to decrease *T* % to 7% (from ∼ 14%).

Finally, another SARS-CoV-2 study reported pooled testing results for 2519 samples performed using a pool size of *n* = 10 or 11.^3^ A total of 1243 tests were required to identify every one of the 241 positive individuals in this population with an average infection rate of *p* = 241*/*2519 = 0.096 (9.6%).^3^ In this case Eq. 4 predicts that a somewhat larger number of 2519 × *T* %(0.096, 10)*/*100 = 1853 tests should have been required to identify every infected sample. The fact that only 1243 rather than 1853 tests were required suggests that a significant number of the positive samples were clustered together in the 99 positive pools, rather than more uniformly distributed over the predicted number of 160 positive pools. However, it is also possible that the differences between the actual and predicted number of positive pools represents a statistical fluctuation associated with the relatively small number of samples tested, as Eq. 4 represents the predicted testing percentage expected when sampling a very large population.

## Summary and Discussion

Optimally pooled testing for SARS-CoV-2 is expected to increases testing efficiency in populations with infection rates below 30%, and becomes more advantageous with decreasing infection rate, as long as the sensitivity of each test is sufficient to detect one infected individual diluted in a pool of optimal size. In a populations with an infection rate of 0.1% the predicted optimal pool size is 32, which is consistent with recently reported SARS-CoV-2 testing sensitivities achievable using a standard RT-qPCR test. ^2,6^ However, in general, a practical upper bound to the pool size is determined by the cycle threshold (*Ct*) pertaining to the population of interest and the particular RT-qPCR test equipment and protocols. ^2,5,11^ Once an acceptable maximum pool size is established, then that maximum pool size should be used instead of the larger optimal pool sizes in Table 1. Moreover, when using a non-optimal first-round pool size, then the optimal second round pool size should be re-calculated using Eqs. 7 and 8. However, when the optimal pool size listed in Table 1 is less than or equal to the maximum pool size, then pooled testing efficiency is expected to improve when using the smaller pool sizes listed in Table 1.

Two rounds of pooled testing are expected to be highly advantageous in facilitating the continuous testing of a populations with a low infection rates. For example, in a population of 100,000 with an average infection probability of 0.1% it is predicted that every infected individual could be identified by performing as few as approximately 4,000 tests, or as few as approximately 15,000 tests in a population of the same size with an infection probability of 1%. This relatively low testing load should make it practical to repeatedly test a population in order to identify early warnings of an emerging outbreak.

Although it is often assumed that pooled testing is only useful for populations with infection rates below about 5%, the present results indicate that even when the infection rate is 10%, pooling can be used to decrease the number of required tests by approximately 40% (relative to individual testing). Moreover, the natural variability of infection rates within a population with a given average infection rate is expected to improve, rather than degrade, the efficiency of pooled testing, as clustering of positive samples in fewer positive pools will decease the number of pools that need to be individually tested.

More generally, optimal pooled testing is expected to be most advantageous when applied to asymptomatic or randomly sampled individuals, as symptomatic individuals are likely to have an infection probability near or exceeding 30%. Thus, pooled testing is expected to be most useful for detection of outbreaks in a relatively stable population, so as to prevent a runaway growth of viral infections. However, controlling such outbreaks requires not only optimal pooled testing but also other factors, including effective isolation and contact tracing.

## Data Availability

The associated Data may be obtained from the author, Dor Ben-Amotz or

## Acknowledgement

This work was supported by the National Science Foundation (Grant Number CHE-109746) and benefited significantly from discussions with Dr. Manoj Jain of the University of Tennessee, Memphis, and Rollins School of Public Health at Emory University in Atlanta.

## Notes

### Competing Interest Statement

The authors have declared no competing interest.

### Funding Statement

NSF Grant Number CHE-109746

### Author Declarations

No IRB/oversight is required for this theoretical predictive study.

